# Prognostic significance of natriuretic peptides after transcatheter aortic valve replacement: A systematic review and meta-analysis

**DOI:** 10.64898/2025.12.02.25341522

**Authors:** Yevhen Kushnir, Nelson Barrera, Erick Romero, Usman Alam, Iurii Statnii, Kristina Golovataya, Maria Camila Tole, Anna E. Bortnick

**Affiliations:** SBH Health System, Department of Internal Medicine, City University of New York School of Medicine, Bronx, NY; Department of Medicine, Division of Cardiology and Division of Geriatrics, Montefiore Medical Center and Albert Einstein College of Medicine, Jack D. Weiler Hospital, Bronx, NY; Yale University School of Medicine, Bridgeport Hospital, Department of Internal Medicine, 267 Grant St, Bridgeport, CT 06610, United States; McLaren Greater Lansing, Department of Internal Medicine, 2900 Collins Rd, Lansing, MI 48910

**Author notes:** Address for correspondence: Yevhen Kushnir, MD; SBH Health System, Department of Internal Medicine, NY 10457, United States. **Sources of funding:** AEB recognizes K23 HL146982 from the National Heart, Lung and Blood Institute (NHLBI), the Resnick Emerging Scholar in Aging Award from the Albert Einstein College of Medicine, and philanthropy from Dr. Hazel J. Chambers. **Disclosures:** AEB served as site principal investigator for multi-center trials sponsored by Amgen, CSL-Behring, and Arrowhead Pharmaceuticals for which her institution received compensation, reports an unrestricted educational grant from Zoll to the institution, honoraria from ClearView HealthCare LLC, and honoraria from Getinge and Zoll to the institution. The remaining authors have nothing to disclose.

**Keywords:** B-type natriuretic peptide, N-terminal pro-beta natriuretic peptide, transcatheter aortic valve replacement, heart failure, mortality, aortic stenosis

## Abstract

**Background:** Elevated B-type- and NT-pro-beta natriuretic peptide (BNP and NT-pro-BNP) after transcatheter aortic valve replacement (TAVR) may be associated with worse outcomes.

**Objective:** We performed a systematic review and meta-analysis to investigate the association of post-TAVR BNP and NT-pro BNP levels on all-cause mortality and heart failure hospitalization.

**Methods:** Seven retrospective studies (n=4143) were included, with a mean follow-up of 32 months. Lower BNP and NT-pro BNP was defined as BNP <202 pg/mL or NT-pro BNP <1423 pg/mL, or BNP that decreased >40% or NT-pro BNP decreased >30% from baseline, or pre/post-TAVR BNP or NT-pro BNP ratio ≥1. Higher BNP and NT-pro BNP was defined as BNP >202 pg/mL or NT-pro BNP >35,000 pg/mL or BNP decreased ≤40% or pre/post-TAVR ratio ≤1 or NT-pro BNP decreased ≤30% from baseline or pre/post-procedure NT-pro BNP ratio < 1.

**Results:** The mean age was 80 ±6 years in the lower group (n=2578) and 81 ±6 years in the higher group (n=1565). Higher BNP and NT-pro BNP persisting after TAVR were associated with a statistically significant increase in all-cause mortality (HR 2.85 [95% CI 2.15-3.78], p<0.00001) and heart failure hospitalization (HR 4.72 [95% CI 2.94-7.57], p<0.00001). A smaller difference between baseline and post-TAVR BNP and NT-pro BNP was associated with a statistically significant increase in all-cause mortality (HR 2.70 [95% CI 1.87-3.92], p<0.00001). Conclusion─Individuals with higher natriuretic peptide levels after TAVR have higher hazards of all-cause mortality and heart failure hospitalization when compared with patients having lower post-procedural levels. Natriuretic peptides measured post-TAVR improve risk stratification.

## Introduction

Aortic stenosis is a prevalent valvular condition in the elderly, leading to left ventricular outflow tract obstruction, heart failure, and increased risk of mortality.(1) Transcatheter aortic valve replacement (TAVR) is a gold-standard treatment for severe symptomatic aortic stenosis in high- and intermediate-risk individuals.(2, 3) Many individuals who undergo TAVR are older with multiple comorbidities and are at risk of periprocedural mortality or heart failure hospitalization. Identifying individuals who would benefit from interventions to decrease mortality and hospitalization following TAVR is imperative.

B-type natriuretic peptide (BNP) and N-terminal pro-beta natriuretic peptide (NT-pro BNP), released from myocytes in response to increased wall stress in severe aortic stenosis, have been studied as prognostic biomarkers in several studies for risk stratification and prediction of clinical outcomes around the time of TAVR.(4) An early study of 31 patients with a 2-month follow-up reported no association between pre-TAVR NT-pro BNP levels and mortality.(5) Similarly, a 2019 meta-analysis found no link between pre-TAVR BNP levels and early mortality (1–2 months) but reported a significant association with midterm mortality (6–48 months).(6)

Several single-center studies have also explored both pre- and post-procedural BNP and NT-pro BNP levels, finding prognostic value when measuring absolute post-TAVR levels or the changes from baseline.(7, 8) However, these studies are limited by small sample sizes, short follow-up durations, and focus on a single outcome.

To address these limitations, we conducted a systematic review and meta-analysis to evaluate (1) the association between post-TAVR BNP or NT-pro BNP levels and all-cause mortality or heart failure hospitalization and (2) the prognostic significance of changes in BNP or NT-pro BNP levels from baseline to post-procedure in predicting all-cause mortality.

## Methods

### Eligibility criteria

Inclusion in this meta-analysis was restricted to studies that met the following eligibility criteria: (1) Randomized trials or non-randomized cohorts; (2) Compared lower vs. higher BNP or NT-pro BNP levels after TAVR or compared higher vs. lower differences between baseline and post-TAVR BNP or NT-pro BNP; (3) BNP or NT-pro BNP was measured after TAVR at least once and/or measured before and after TAVR; and (4) follow-up for 12 months or more.

“Lower” post-TAVR BNP and NT-pro BNP was defined as BNP <202 pg/mL or NT-pro BNP <1423 pg/mL, or BNP levels that decreased >40% from baseline, or NT-pro BNP levels that decreased >30% from baseline, or pre/post-TAVR BNP or NT-pro BNP ratio ≥1. “Higher” post-TAVR BNP and NT-pro BNP was defined as BNP >202 pg/mL or NT-pro BNP >35,000 pg/mL or BNP level that decreased ≤ 40% from baseline or pre/post-TAVR ratio ≤1 or NT-pro BNP level decreased ≤ 30% from baseline or pre/post-procedure NT-pro BNP ratio < 1.We excluded studies with no BNP or NT-pro BNP level measured after TAVR.

### Search strategy and data extraction

We systematically searched PubMed, Scopus, and the Cochrane Central Register of Controlled Trials from inception to December 2024 using the search term “BNP OR NT-pro BNP AND TAVR” We also manually searched the references from all included studies for additional studies. Two authors (Y.K and N.B) independently extracted the data after predefined search criteria and quality assessment. The meta-analysis protocol was registered on PROSPERO, 2024, under protocol <u>#CRD42024621496</u>.

### Endpoints and sensitivity analysis

The outcomes of interest were all-cause mortality and heart failure hospitalization. We performed a leave-one-out sensitivity analysis to investigate the impact of excluding individual studies on the cumulative analysis for all-cause mortality and heart failure.

### Quality assessment

We evaluated the risk of bias in non-randomized studies using the Risk of Bias in Non-randomized Studies—of Interventions tool (ROBINS-I).(9) Two independent authors (Y.K. and N.B) completed the risk of bias assessment (Supplementary Table 1). Disagreements were resolved through consensus after discussing the reasons for the discrepancy. Publication bias was investigated using funnel-plot analysis of point estimates about study weights. The funnel plot analysis revealed that studies had a symmetrical distribution according to weight and converged toward the pool effect as the weight increased (Fig. S5).

### Statistical analysis

This systematic review and meta-analysis were performed and reported in accordance with the Cochrane Collaboration Handbook for Systematic Reviews of Interventions and the Preferred Reporting Items for Systematic Reviews and Meta-Analysis (PRISMA) Statement guide.(10) Hazard ratios (HR) with 95% confidence intervals were used to compare treatment effects for categorical endpoints. We assessed heterogeneity with I² statistics and the Cochran Q test; p values <0.10 and I² >25% were considered significant for heterogeneity. We used the DerSimonian and Laird random-effects models and Review Manager (Cochrane Center, the Cochrane Collaboration, Denmark) for statistical analysis. P <0.05 was considered statistically significant. We performed meta-regression analyses using the metafor package in R (version 4.5.2; R Foundation for Statistical Computing, Vienna, Austria). Univariable and multivariable random-effects meta-regressions were conducted with the restricted maximum likelihood (REML) estimator. Moderators included mean age, follow-up duration, left ventricular ejection fraction, female proportion, and prevalence of prior coronary revascularization (CABG and PCI). Regression coefficients (β) and corresponding Hartung–Knapp–Sidik–Jonkman (HKSJ) adjusted p-values were reported. Variables were analyzed as continuous covariates, expressed per unit or 10% increase as appropriate. Meta-regression for NYHA functional class was not performed because of inconsistent reporting (some studies reported class III, others class IV, or combined categories). All analyses were two-tailed with p < 0.05 considered statistically significant.

## Results

### Study Selection and Baseline Characteristics

The initial search yielded 136 results. After removing duplicates and ineligible studies, 20 remained and were reviewed for inclusion criteria (Fig. 1). Of those, 7 studies were included in qualitative and quantitative review after the exclusion of 7 with BNP/NT-pro BNP measured only before TAVR. N=4143 were included from 7 retrospective cohorts. N=2578 were included in the lower BNP and NT-pro BNP group, and n=1565 were included in the higher BNP and NT-pro BNP group. Baseline characteristics are shown in Table 1. The mean age was comparable between groups; 80 ± 6 years in the lower and 81 ± 6 years in the higher BNP and NT-pro BNP group. The low BNP/NT-pro BNP group had slightly more favorable baseline characteristics, including higher mean ejection fraction and lower rates of prior coronary artery bypass grafting, prior percutaneous coronary intervention, and NYHA Class III/IV heart failure. The mean follow-up duration across studies was 32 months.

**Figure 1.**
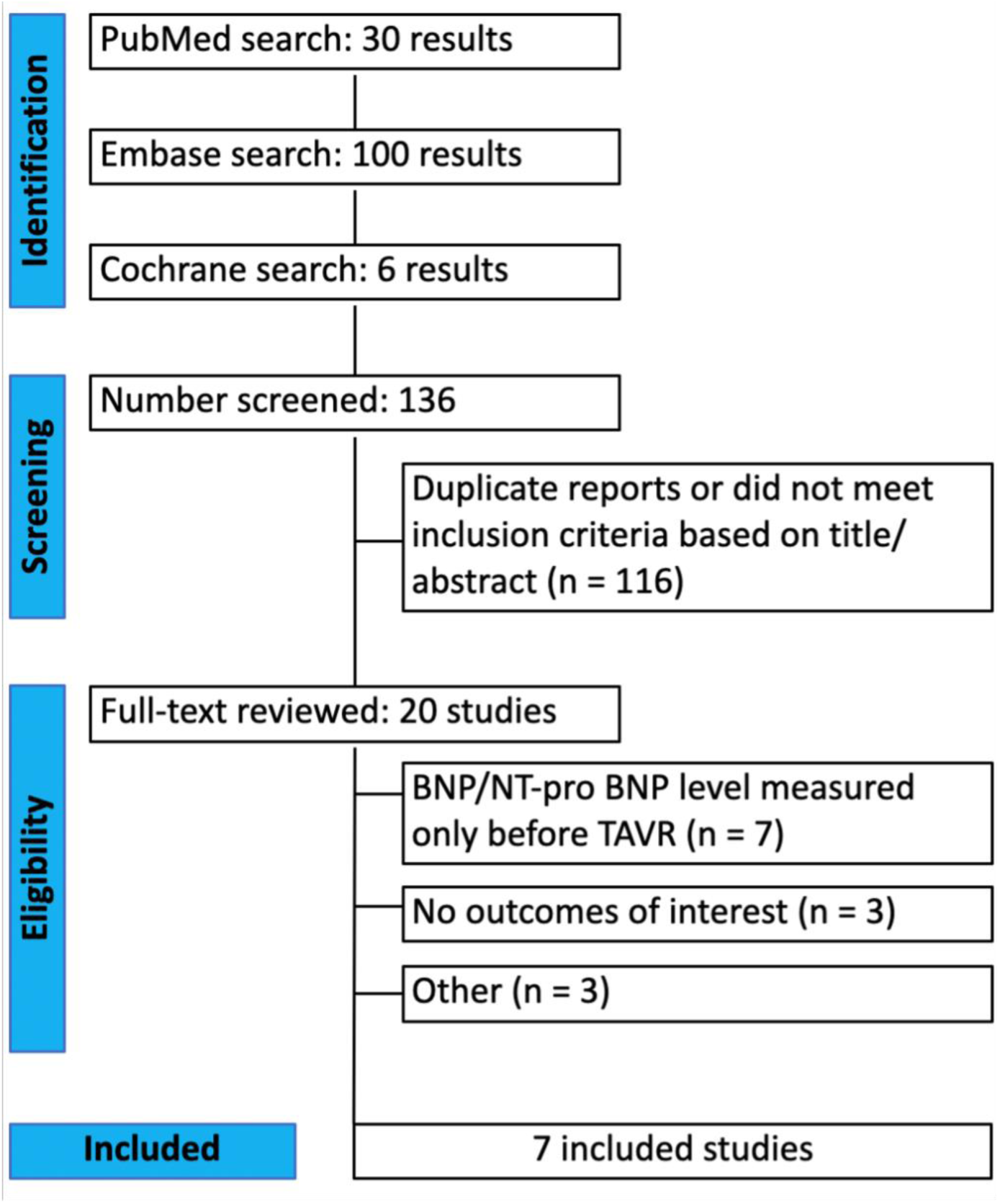
PRISMA flow diagram of study screening and selection. BNP=B-type natriuretic peptide, NT-pro BNP =N-terminal pro-beta natriuretic peptide, TAVR=transcatheter aortic valve replacement.

**Table 1.** Baseline characteristics of included studies, comparing lower and higher BNP and NT-pro BNP groups. + - BNP ratio was calculated as the measured circulating BNP level divided by the upper limit of normal for the BNP. CABG – coronary artery bypass graft, EF – ejection fraction, NP - natriuretic peptides, NYHA – New York Heart Association heart failure classification PCI – percutaneous coronary intervention

| Study | Type of NP* | Center | Low/high NP*criteria | When NP* was measured | n of patients low/high NP* | Months of follow up | Age, mean low/high | Female, % low/high | EF, % low/high | Prior CABG, % low/high | Prior PCI,% low/high | NYHA III and/or IV, % low/high |
| --- | --- | --- | --- | --- | --- | --- | --- | --- | --- | --- | --- | --- |
| O' Leary(16) | BNP | 4; US | Q1 BNP ratio tertile(136)/Q4 BNP ratio tertile(614)+ | Baseline, discharge, 30 days, 1 year | 534/532 | 24 | 80.7 ± 7.5/82.7± 7 | 47.9/42.3 | 61.8/52.5 | 23.4/29.1 | 26/33.1 | 82.2/77.6 |
| Yu(7) | NT-pro BNP | 1; China | NT-proBNP ratio ≥ 1/NT-proBNP ratio < 1 | 48h before TAVR, 4-8 days after TAVR | 234/123 | 53 | 71.9± 8.5/73.3± 6.3 | 41/46 | 63/58 | 3.0/4.1 | 12.8/17.1 | 68.4/74.0 |
| Kaneko(15) | NT-pro BNP | 1; Germany | NT-proBNP levels decreased >30% from baseline/≤ 30% from baseline | Baseline, at discharge | 448/269 | 24 | 80 ± 7/81 ± 7 | 50/59 | 45/50 | 11/13 | 36/46 | 16/16 (IV only) |
| Mizutani(17) | BNP | 14; Japan | BNP ≤202 pg/mL/BNP >202 pg/mL | At discharge after TAVR | 576/518 | 24 | 85(81–87)/86(82–88) | 69.4/72.2 | 63.8/60 | 6.9/6.9 | N/A | 3.5/9.1 (IV only) |
| Onoda(8) | BNP | 1; Japan | BNP levels decreased >40% from baseline/≤ 40% from baseline | Before TAVR/1-2 weeks after TAVR | 48/22 | 16.3 ± 9 | 86 ± 4./85 ± 5 | 79.2/72.7 | 57.6/59.3 | N/A | N/A | 34/12 |
| Phan(18) | BNP | 1; US | Pre/post TAVR ratio >1/ Pre/post TAVR BNP ratio ≤ 1) | Pre-TAVR, 6-12 months after TAVR | 77/78 | 23 | 82 ± 7 | 49 | N/A | N/A | N/A | N/A |
| Zhou(14) | NT-pro | 1; China | 1423 pg/mL/35000 | Baseline, discharge, | 661/33 | 60 | 74± 5/76± 5.5 | 43.42/40 | 60.5/42 | 0.3/0 | 11.3/31.4 | 25.42/48.57 |
|  | BNP |  | pg/mL | 30 days |  |  |  |  |  |  |  | (IV only) |
| Mean values | N/A | N/A | N/A | N/A | N/A | 32 | 80 ±6 years/81 ±6 years | 54.2/54.4 | 58.5/53.6 | 7.4/8.9 | 21.5/31.9 | 38.2/39.5 |

### Pooled Analysis

Individuals with higher BNP and NT-pro BNP levels post-TAVR had statistically significant higher hazards of all-cause mortality compared to individuals with lower BNP and NT-pro BNP levels (HR 2.85 [95% CI 2.15-3.78], p<0.00001, Fig. 2A, Central Illustration). The results showed a low heterogeneity with I^2^=6% (Fig. 2A). Similarly, individuals with higher BNP and NT-pro BNP levels post-TAVR had statistically significant higher hazards of heart failure hospitalization compared to individuals with lower BNP and NT-pro BNP level post-TAVR (HR 4.72 [95% CI 2.94-7.57], p<0.00001, Fig. 2B, Central Illustration). The results showed no heterogeneity with I^2^=0% (Fig. 2B). Lastly, individuals with a smaller reduction between baseline and post-TAVR BNP or NT-pro BNP levels had statistically significant higher hazards of all-cause mortality compared to those with a greater reduction between baseline and post-TAVR BNP or NT-pro BNP levels (HR 2.70 [95% CI 1.87-3.92], p<0.00001, Fig 3). The results showed low heterogeneity with I^2^=0% (Fig 3).

**Figure 2A.**
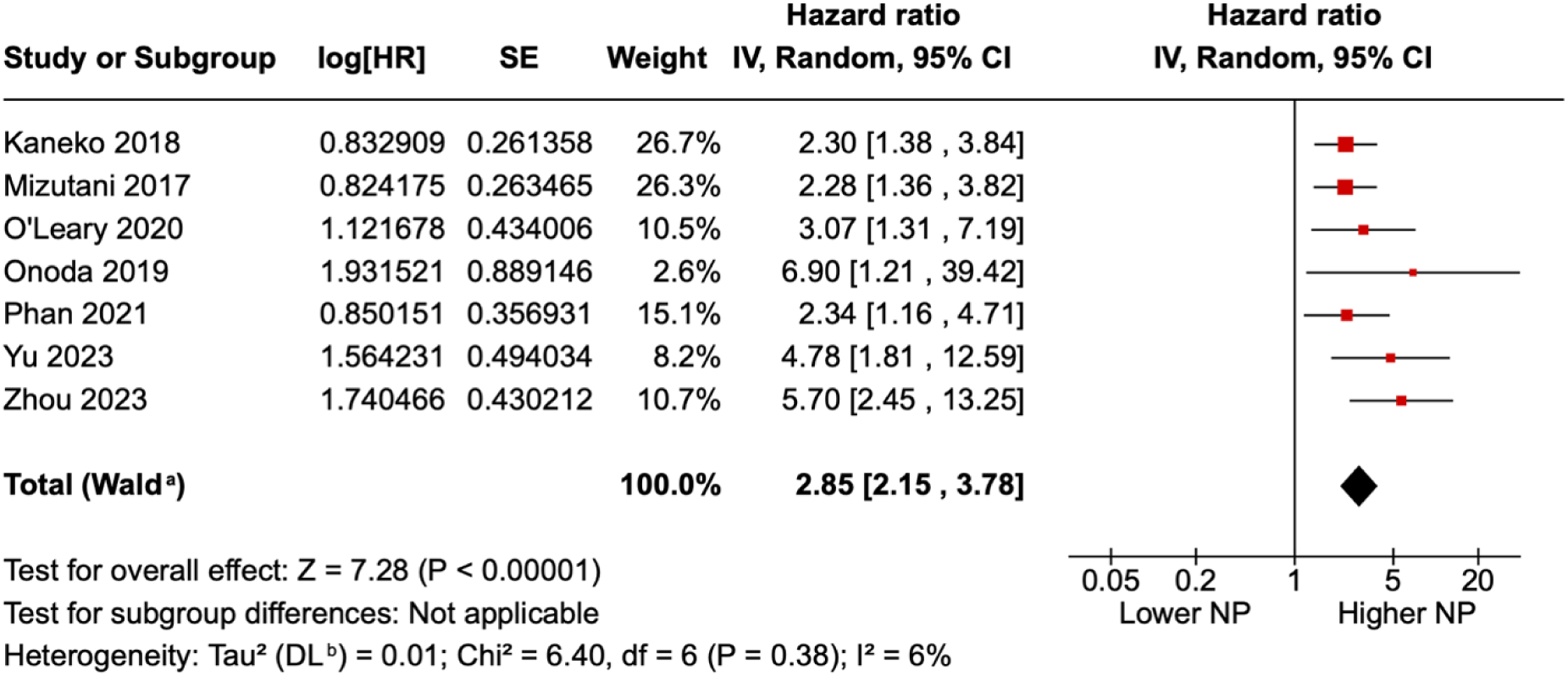
Forest plot showing the hazards of all-cause mortality in lower and higher BNP and NT-pro BNP groups. The definition of lower and higher BNP/NT-pro BNP level varies through the studies: in some studies it was descriptive number (Mizutani: lower BNP group - BNP ≤202 pg/mL, higher BNP group - BNP >202 pg/mL), in some studies it was level difference before and after TAVR (Onoda: lower BNP group BNP levels decreased >40% from baseline, higher BNP group - ≤ 40% from baseline). HR=hazard ratio, SE=standard error. BNP=B-type natriuretic peptide, NT-pro BNP =N-terminal pro-beta natriuretic peptide. NP=natriuretic peptides.

**Figure 2B.**
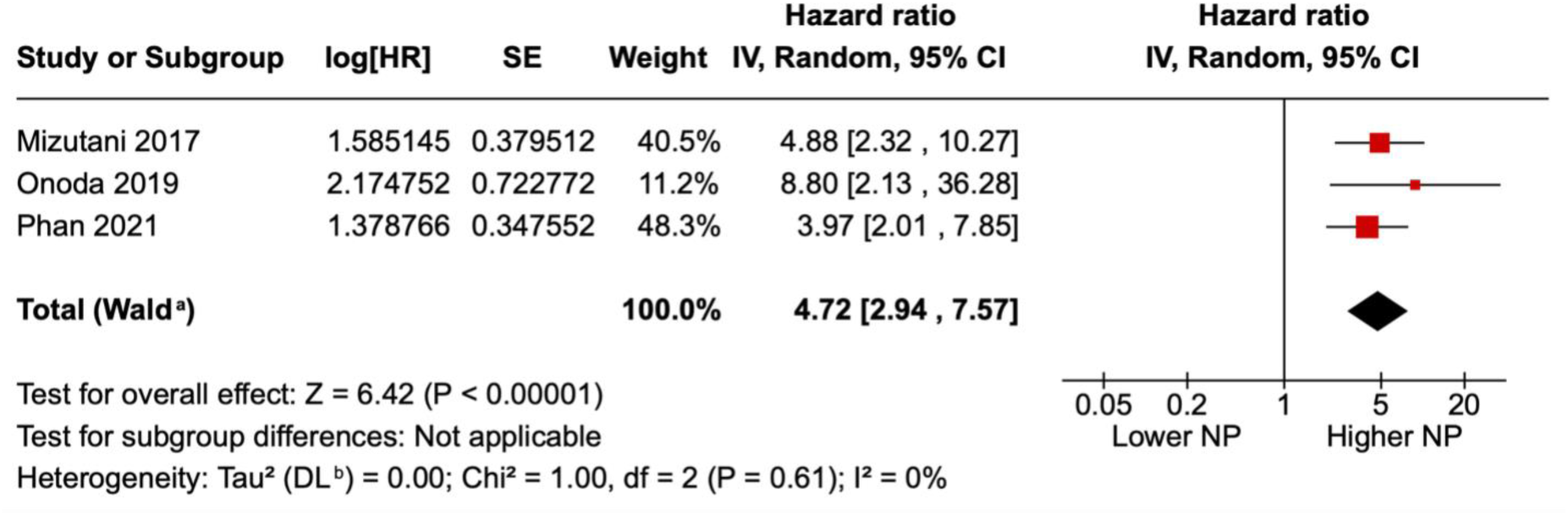
Forest plot showing the hazards of heart failure hospitalization in lower and higher BNP and NT-pro BNP groups. The definition of lower and higher BNP/NT-pro BNP levels varies through the studies: in some studies it was defined level (Mizutani: lower BNP group - BNP ≤202 pg/mL, higher BNP group - BNP >202 pg/mL), in some studies it was level difference before and after TAVR (Onoda: lower BNP group - BNP levels decreased >40% from baseline, higher BNP group - BNP levels decreased ≤ 40% from baseline). HR=hazard ratio, SE=standard error. BNP=B-type natriuretic peptide, NT-pro BNP =N-terminal pro-beta natriuretic peptide. NP=natriuretic peptides.

**Figure 3.**
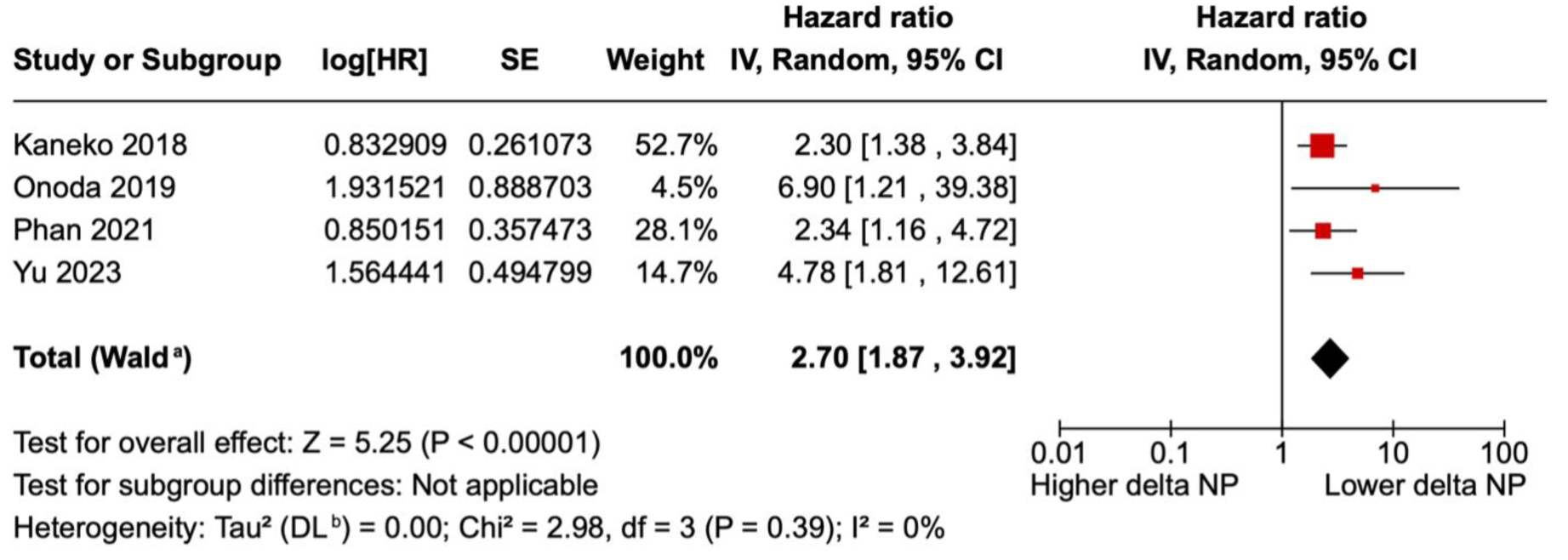
Forest plot showing the hazards of all-cause mortality in lower and higher differences between baseline and post-TAVR BNP and NT-pro BNP groups. Delta NP: the difference between baseline and post-TAVR BNP and NT-pro BNP. HR=hazard ratio, SE=standard error. BNP=B-type natriuretic peptide. NT-pro BNP =N-terminal pro-beta natriuretic peptide, NP=natriuretic peptides.

### Meta-regression

Meta-regression identified follow-up duration as a significant moderator (β = 0.0216 ± 0.0073; p = 0.029), indicating that longer follow-up periods strengthened the observed association. No significant modifying effects were found for mean age, left ventricular ejection fraction, or female proportion. The multivariable model including both age and ejection fraction was non-significant (p = 0.19).

In univariable analyses, the proportion of patients with prior CABG did not influence the mortality association (k = 5; β = −0.018 [SE 0.024]; HKSJ p = 0.50; OR per 10% increase ≈ 0.83, 95% CI crossing 1). Conversely, a higher prevalence of prior PCI was associated with a weaker relationship between natriuretic peptide levels and mortality (k = 4; β = −0.031 [SE 0.0015]; HKSJ p = 0.0024), suggesting approximately 26% lower odds ratio per 10% increase in prior PCI. However, this finding should be considered exploratory given the limited reporting (k ≤ 5). The detailed results of the meta-regression analyses are presented in **Table 2**. Meta-regression for NYHA functional class was not performed because of inconsistent reporting—some studies provided data solely for class IV, others for class III, and a few combined both, precluding harmonized quantitative analysis.

**Table 2.** Univariable and Multivariable Meta-Regression Analyses for All-Cause Mortality. Meta-regression analyses were performed to explore potential moderators of the association between elevated natriuretic peptide levels and all-cause mortality. The regression coefficient (β) represents the change in log odds ratio per unit increase in each moderator. Positive β values indicate stronger associations, whereas negative β values suggest attenuation. p-values were calculated using the Hartung–Knapp–Sidik–Jonkman (HKSJ) adjustment under a random-effects model with the Restricted Maximum-Likelihood (REML) estimator. Follow-up duration emerged as a significant moderator, indicating stronger mortality associations with longer observation periods. No significant effects were observed for mean age, ejection fraction, or female proportion. Among procedural variables, prior CABG showed no influence, while a higher prevalence of prior PCI was associated with a weaker relationship between natriuretic peptide levels and mortality. The NYHA functional class variable was excluded due to inconsistent reporting across studies. Notes: β represents the slope of the meta-regression (change in log OR per unit increase in moderator). HKSJ = Hartung–Knapp–Sidik–Jonkman adjustment. All analyses used a random-effects model with the REML estimator. NYHA meta-regression was not performed because of heterogeneous reporting (class III only, class IV only, or combined III–IV categories).

| Moderator Variable | k (studies) | Model Type | $\beta$ (Coefficient ± SE) | HKSJ p-value | Direction / Interpretation |
| --- | --- | --- | --- | --- | --- |
| Follow-up duration (months) | 7 | Univariable | $0.0216 \pm 0.0073$ | 0.029 | Longer follow-up associated with stronger mortality association |
| Mean age (years) | 7 | Univariable | — | >0.05 | No significant effect |
| Left ventricular ejection fraction (%) | 7 | Univariable | — | >0.05 | No significant effect |
| Female proportion (%) | 7 | Univariable | — | >0.05 | No significant effect |
| Age + Ejection Fraction | 7 | Multivariable | — | 0.19 | Combined model non-significant |
| Prior CABG (%) | 5 | Univariable | $-0.018 \pm 0.024$ | 0.50 | No modification of mortality association (OR per 10% $\uparrow \approx 0.83$) |
| Prior PCI (%) | 4 | Univariable | $-0.031 \pm 0.0015$ | 0.0024 | Higher PCI prevalence associated with weaker mortality association (~26% lower OR per 10% ↑) |
| NYHA III/IV (%) | — | — | — | — | Not analyzed due to inconsistent reporting across studies |

### Sensitivity analysis

Individuals with higher NT-pro BNP levels post-TAVR had statistically significant higher hazard of all-cause mortality compared to individuals with lower post-TAVR levels (HR 3.62 [95% CI 1.94-6.73], p<0.0001, Fig. S1). The results showed moderate heterogeneity with I^2^=51% (Fig. S1). Individuals with higher BNP levels post-TAVR had a statistically significant higher hazard of all-cause mortality compared to those with lower levels (HR 2.55 [95% CI 1.67-3.67], p<0.00001, Fig. S2). The results showed a low heterogeneity with I^2^=0% (Fig S2).

Leave-one-out sensitivity analysis was performed to explore the impact of separate studies on the cumulative analysis for all-cause mortality (Fig. S3) and heart failure hospitalization (Fig. S4). The results were similar to the primary analysis.

## Discussion

We investigated the association between post-TAVR BNP and NT-pro BNP levels on all-cause mortality and heart failure hospitalization, as well as differences between baseline and post-TAVR BNP and NT-pro BNP levels on all-cause mortality in a meta-analysis of 7 studies with n=4143 individuals. Individuals with higher BNP or NT-pro BNP levels experienced a 2.85-fold increase in the hazard of all-cause mortality and a 4.72-fold increase in the hazard of heart failure hospitalization after the procedure. Persistently higher post-TAVR BNP or NT-pro BNP levels were associated with a 3.62-fold increase in the hazard of all-cause mortality and a 2.55-fold increase in the hazard of all-cause mortality. Lower difference between baseline and post-TAVR BNP and NT-pro BNP levels was associated with 2.7-fold increase in the hazard of all-cause mortality. This study extends previous research by providing a comprehensive analysis of outcomes in patients with higher and lower post-TAVR BNP and NT-pro BNP levels, resolving the limitations of earlier studies.(6) These data advance BNP and NT-pro BNP as useful biomarkers of TAVR-related all-cause mortality and HF hospitalization. The JAHA 2025 study similarly identified baseline NT-pro BNP as a predictor of long-term mortality following TAVI, aligning with our pooled findings.(11) However, 30-day biomarker changes were not associated with two-year outcomes, and the absence of quantitative effect measures precluded inclusion in the meta-analysis. This underscores that baseline natriuretic peptide burden may better capture long-term risk than short-term fluctuations influenced by peri-procedural factors. Our meta-regression analysis identified follow-up duration as a significant moderator, indicating that studies with longer follow-up periods demonstrated stronger associations between elevated natriuretic peptide levels and all-cause mortality. In contrast, demographic and clinical variables such as age, ejection fraction, and female proportion did not significantly influence the effect size. These findings suggest that the prognostic impact of natriuretic peptides becomes more pronounced over time, likely reflecting the cumulative progression of ventricular dysfunction and adverse remodeling.

Elevated levels of BNP and NT-pro BNP are routinely used as biomarkers for the diagnosis of HF.(12) BNP and NT-pro BNP levels measured prior to TAVR for risk stratification and prediction of clinical outcomes has demonstrated conflicting results. An earlier study of 31 patients with 2-month follow-up reported no association between pre-TAVR NT-pro BNP level and mortality.(5) A 2019 meta-analysis(6) demonstrated lack of an early association with all-cause mortality, but did find an association with increased middle term all-cause mortality at 6-48 months. Various single and multicenter studies of change in BNP and NT-pro BNP levels before and after TAVR, or residual post-TAVR BNP and NT-pro BNP levels demonstrated that smaller change or higher post TAVR levels were associated with clinical outcomes(7) such as all-cause mortality and heart failure hospitalization. Nevertheless, limitations of these studies, like small cohort size and short-term follow-up, hamper generalizability.(8) Individuals with a higher difference between baseline and post-TAVR BNP or NT-pro BNP tended to have more comorbidities with a higher transvalvular velocity and more CHF.(7) Heart failure is the most common cause of hospitalization in patients with TAVR.(13) Those at higher risk of heart failure hospitalization might benefit from more intensive management, adding sodium-glucose cotransporter-2 (SGTL-2) inhibitors or adding advanced heart failure specialists to the care team. Another approach to management would be to serially measure BNP or NT-pro BNP levels following TAVR and follow those with a steeper upward trajectory more closely.(14)

## Strengths and limitations

This study had several strengths. It is the first meta-analysis evaluating the association of post-TAVR BNP and NT-pro BNP levels and the difference between baseline and post-TAVR BNP and NT-pro BNP with long-term mortality and heart failure hospitalization. However, we acknowledge several limitations. First, the analysis included non-randomized studies. Second, there was variability in the definitions and ranges of lower and higher BNP/NT-pro BNP groups, and thus we created a composite definition. We included single and multicenter studies from the US, Europe and Asia, therefore, there may be variability due to differences in laboratory assays, but generalizability was improved by including a diverse population. CKD and use of a neprilysin inhibitor (sacubitril-valsartan) may have been confounders and were not controlled for. BNP and NT-pro BNP clearance decreases in CKD and only 2 of 7 studies included in the analysis reported the prevalence of CKD. In one study, there was no statistically significant difference in CKD prevalence between the groups.(7) In another study, there was a significant difference in the prevalence of CKD between groups (51% in responders and 66% in non-responders group), CKD was an independent predictor of NT-pro BNP nonresponse, however CKD itself was not directly listed as an independent predictor of mortality in the Cox regression.(15) Neprilysin inhibition decreases clearance of BNP and none of the 7 studies provided information on sacubitril-valsartan use. Due to the limited number of studies reporting heart failure hospitalization as an outcome, direct comparisons between BNP and NT-pro BNP, as well as difference between post-TAVR levels and changes from baseline, could not be performed. Older adults with a mean age of ≥80 years most commonly underwent TAVR in these studies, therefore findings are not generalizable to a younger population.

## Conclusions

In this meta-analysis, we found that higher natriuretic peptide levels in patients after TAVR were associated with increased hazards of all-cause mortality and heart failure hospitalization when compared to those with lower levels. Both markers (BNP vs. NT-pro BNP) and approaches (post-TAVR or the difference between baseline and post-TAVR) was associated with all-cause mortality and heart failure hospitalization. Further studies implementing risk stratification and treatment are warranted.

## Data Availability

All data supporting the findings of this study are available within the article and its online supplementary materials. The extracted datasets used for the meta-analysis were derived from previously published studies, which are publicly available through their respective journals. No new patient-level data were collected.

## Abbreviations

CI: Confidence interval
HR: hazard ratio
PRISMA: Preferred reporting items for systematic reviews and meta-analysis
PROSPERO: International Prospective Register of Systematic Reviews
ROBINS – I: Risk of Bias in Non-randomized Studies of interventions
BNP: B-type natriuretic peptide
NT-pro BNP: N-terminal pro-beta natriuretic peptide
TAVR: transcatheter aortic valve replacement
HF: heart failure
EF: ejection fraction

## Clinical perspectives

Future studies should use natriuretic peptide levels after TAVR to risk-stratify management strategies to decrease long-term all-cause mortality and heart failure hospitalization.

## Highlights

### What is known?

1) Higher post-TAVR and 2) lower delta BNP and NT-pro BNP are associated with higher all-cause mortality.

### What is new?

Both markers (BNP/NT-pro BNP) and approaches (post-TAVR/delta) were associated with all-cause mortality.

### What is next?

To prospectively test the effectiveness of post-TAVR/delta natriuretic peptide levels for risk stratification.

## Acknowledgments

We appreciate the valuable review and insightful comments by Dr. Sandhya Murthy, Montefiore Medical Center and Albert Einstein College of Medicine, Bronx NY. We appreciate the statistical review by Dr. Ryung Kim, Albert Einstein College of Medicine, Bronx NY.

## Data Sharing

Data analysis available upon request.

## Ethics

This systematic review and meta-analysis used data from published studies, which are publicly available. An aggregated data from individual studies that do not include identifiable details of study participants. The study was performed and reported in accordance with the Cochrane Collaboration Handbook for Systematic Reviews of Interventions and the Preferred Reporting Items for Systematic Reviews and Meta-Analysis (PRISMA) Statement guide. The meta-analysis protocol was registered on PROSPERO, 2024, under protocol #CRD42024621496.

## Central illustration

Seven retrospective studies of 4143 patients with a mean age of 81 years and a mean follow-up period of 32 months, where 2578 were included in the lower BNP and NT-pro BNP group and 1565 were included in the higher BNP and NT-pro BNP group. Pooled analysis showed higher odds of all-cause mortality (HR 2.85) and heart failure hospitalization (HR 4.72) in the higher BNP and NT-pro BNP group when compared to the lower BNP and NT-pro BNP group.

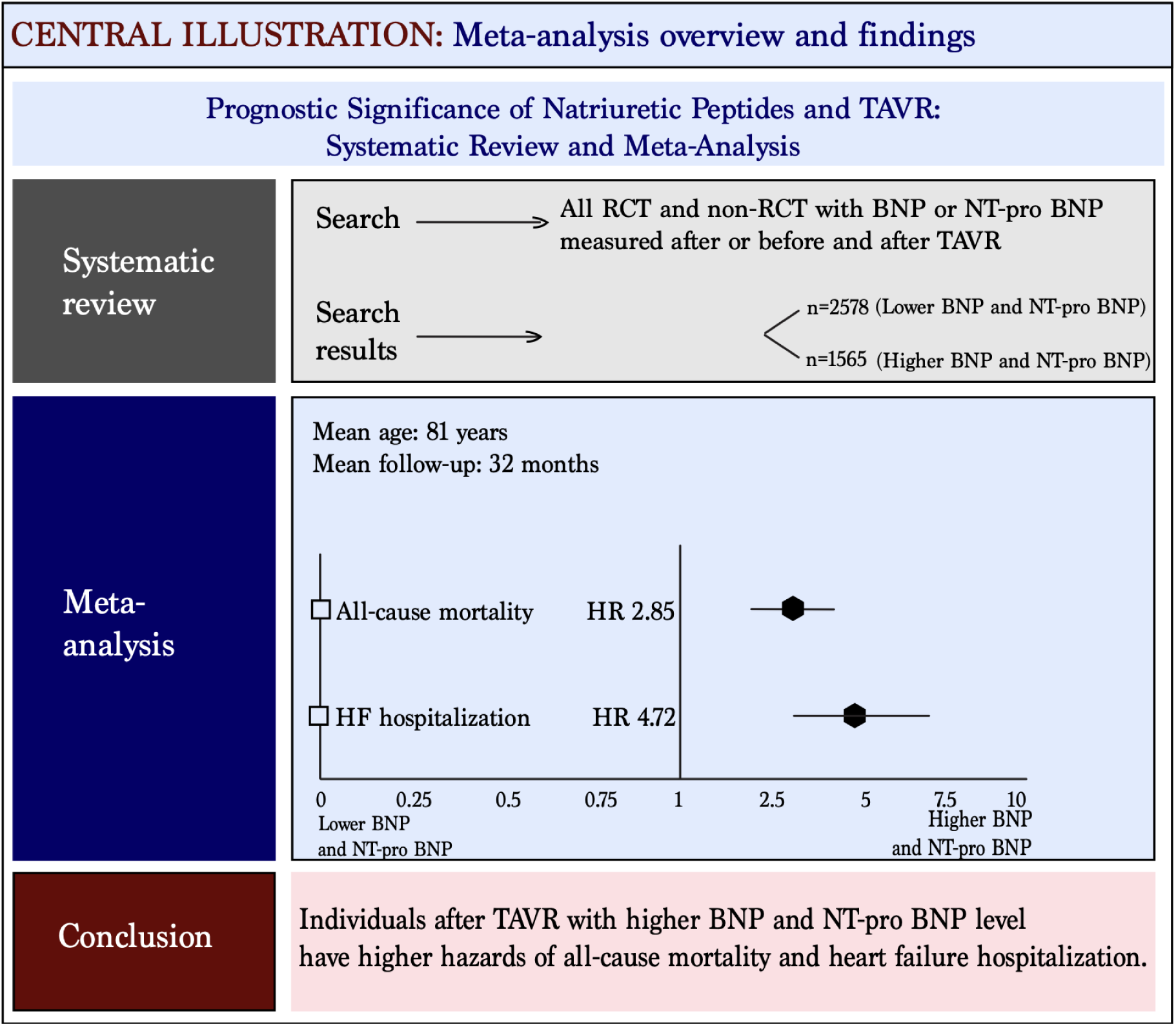

